# FoxTail: An R-Peak-Anchored Event Domain for Visualizing and Quantifying Changes in ECG Dynamics

**DOI:** 10.64898/2026.08.16.26360545

**Authors:** Nuno M. Garcia

## Abstract

Conventional electrocardiography is highly effective for waveform and rhythm diagnosis, but it is less suited to showing how the internal shape of hundreds or thousands of consecutive heartbeats changes over time. We introduce FoxTail, a complementary view that represents each cardiac cycle as an ordered sequence of changes in signal direction. Overlaying these sequences in a fixed visual field makes beat-to-beat organization visible and allows the density, size, stability, and scale persistence of those changes to be measured. We evaluated the representation in recordings containing normal sinus rhythm, paroxysmal atrial fibrillation, severe heart failure, ventricular tachyarrhythmia, and controlled electrode-motion noise. Paired recordings showed that FoxTail descriptors can reveal within-person state changes that are not conveyed by a single average beat. The noise and pre-fibrillation analyses also showed that a dense event pattern is not automatically equivalent to physiological complexity, measurement artifact, or impending disease. FoxTail is therefore not proposed as a replacement for the diagnostic ECG or as a new classifier, but as an observation and measurement domain for asking a more basic question: how is the electrical organization of the heart changing from one beat to the next, and which of those changes persist across scale?

## I. Introduction

The electrocardiogram has been extraordinarily successful because it makes cardiac electrical activity visible. Its conventional amplitude-versus-time representation supports rhythm identification, interval measurement, waveform interpretation, and diagnosis. The problem addressed here is not a failure of that representation, but a different observational need: a long recording contains the evolving relationship between successive cardiac cycles, and the way in which their internal trajectories differ is difficult to perceive in a conventional display.

A monitor solves the problem of temporal continuity by scrolling, but only a few cycles remain visible at useful morphological resolution; a long printed strip enlarges the visual field without changing this limitation; ensemble averaging solves a different problem by constructing a representative beat, but variation between beats is treated as dispersion around that representative and is consequently attenuated.

Heart-rate variability and nonlinear analyses quantify important aspects of cardiac dynamics, yet they commonly operate on R–R intervals or summary features rather than on the ordered within-cycle changes of the ECG waveform [1]– [3]. There remains a representational gap between inspecting individual morphologies and reducing a long recording to aggregate descriptors.

FoxTail draws conceptual inspiration from the Eye Diagram [4], [5] used in optical communications. An Eye Diagram superimposes many symbol intervals within a common reference frame, allowing timing jitter, amplitude noise, distortion, and other departures from the expected transition pattern to become visible as an ensemble structure. The analogy is not exact, as cardiac cycles are neither identical symbols nor fixed-duration intervals, but it suggests a powerful observational strategy: align many cycles to a physiologically meaningful landmark and allow their differences to coexist within the same visual field. In an ECG, those differences are not merely unwanted deviations because they may express the changing electrical dynamics that make the signal physiologically informative. Even so, canonical ECG representations don’t allow the assessment of the changes in the dynamics that occur from heartbeat to heartbeat.

FoxTail is proposed to fill this gap as an exploratory and longitudinal observation domain for comparing states within an individual, studying differences between individuals under controlled acquisition, and locating intervals in which cardiac dynamics change. Clinical classifiers may eventually be built on this domain, but diagnosis is neither assumed nor required for the representation itself to be informative.

Within each R-anchored cardiac cycle, FoxTail distinguishes between samples that continue an existing monotonic rise or fall and samples that reverse the direction of the trajectory. Samples in the first group refine the path between successive extrema, whereas samples in the second define new directional events. This distinction is algorithmic rather than physiological as an omitted sample may still contain clinically relevant information, and a retained turning vertex is not inherently pathological. The question addressed by FoxTail is whether representing each cycle as an ordered sequence of directional events, while retaining amplitude excursions, omitted-run lengths, R—R duration, and original sample positions as associated information, makes changes in ECG dynamics easier to perceive and quantify.

To answer that question, FoxTail changes the observational domain. The signal is divided into natural, variable-duration R-to-R cycles, after which samples that only continue a monotonic run are suppressed and the run endpoints are retained. The resulting sequences are overlaid from a common R anchor and indexed by event order rather than elapsed time. Their image resembles a fox tail, with a dense proximal region formed by recurrent early events and a variable distal region formed by cycles containing progressively more turning vertices. Absolute retained amplitudes provide an immediately interpretable visual form, while amplitude differentials between consecutive vertices provide the principal analytical form.

The central contribution is therefore the change of observational domain, not the preservation of a particular scalar property. Although exact preservation of discrete total variation provides a useful mathematical guarantee, the purpose of the method is to make changes in ECG organization perceptible and measurable across beats and scales. FoxTail is likewise not proposed as a diagnostic classifier; its primary hypothesis is that the event-domain organization of the ECG contains information about changing physiological state that complements conventional morphology and timing.

We test this hypothesis as a measurement problem rather than as a classification contest. The evaluation first establishes what the representation looks like in normal sinus rhythm, severe heart failure, paroxysmal atrial fibrillation recordings, and ventricular tachyarrhythmia. It then asks whether paired recordings reveal within-subject changes around PAF, determines how controlled electrode-motion noise alters the same descriptors, and examines whether a common short-term transition appears before annotated ventricular fibrillation. Taken together, these experiments are intended to distinguish a visually interesting transformation from a scientifically informative observation domain.

The remainder of this paper is organized as follows. Section II positions FoxTail within the literature on ECG morphology, cardiac dynamics, multiscale complexity, and event-based signal representations. Section III presents the mathematical formulation of the FoxTail transformation and defines its principal descriptors. Section IV describes the databases, signal-processing pipeline, experimental questions, and statistical methods. Section V presents the visual and quantitative results, while Section VI discusses their interpretation, limitations, and implications for future validation. Section VII concludes the paper.

## II. From ECG Morphology to Signal Dynamics: Background and Related Work

### A. Morphology, Timing, and Beat-Level Comparison

The conventional ECG is an amplitude-versus-time representation, and its clinical value depends precisely on the fact that it preserves morphology and timing. The P wave, QRS complex, ST segment, and T wave correspond to physiologically meaningful phases of cardiac activation and recovery, while their amplitudes, durations, and relative positions support rhythm identification and diagnosis. Rather than replacing this view, FoxTail begins from a limitation that becomes increasingly apparent as the duration of a recording grows: the conventional display is well suited to examining a small number of cycles in detail, but it is less effective at showing how their internal trajectories vary across hundreds or thousands of beats.

Several established methods extend conventional inspection beyond a single cycle. Beat templates, signal-averaged ECGs, and morphology clustering compare cycles after temporal registration, phase normalization, or averaging. These operations are valuable because they reveal representative morphology and reduce incidental variation. However, that same reduction can make beat-to-beat variation less visible, especially when the variation itself, rather than the nominal waveform, is the object of interest. A long recording therefore creates a representational problem: one may inspect individual beats or summarize the recording, yet it remains difficult to observe how within-cycle organization changes continuously over time.

### B. Cardiac Dynamics and Multiscale Complexity

Cardiac variability is not simply unstructured fluctuation around a nominal rhythm. Analyses of heartbeat dynamics have shown organization across multiple temporal scales, including scale-dependent, fractal, and multifractal behavior. Studies of interbeat intervals demonstrated that this organization changes with aging and disease, suggesting that physiological information lies not only in the average rate or the magnitude of variability, but also in the way fluctuations are arranged across scales [1], [2], [6], [7].

This perspective motivated entropy-based, fractal, recurrence, and multifractal measures of cardiac dynamics. Multiscale entropy made a particularly important distinction between irregularity and complexity: a signal can fluctuate rapidly and remain structurally simple, whereas physiological complexity implies organized behavior that persists across scales [3], [8]. The distinction is directly relevant to ECG analysis because measurement noise may create numerous local reversals without adding meaningful physiological organization. Any event-domain representation must therefore examine event density together with amplitude, temporal stability, and persistence across scale rather than treating a large number of events as evidence of complexity.

Most foundational work in this area has focused on R–R interval series. That perspective captures the timing of cardiac cycles but not the ordered electrical trajectory within each cycle. Multifractal analyses applied directly to ECG waveforms suggest that scale-dependent structure is also present in the morphology of the sampled signal [9]. These findings motivate a representation that retains R–R timing while making within-cycle changes available for visual and quantitative study. They do not, however, imply that every ECG-derived quantity is fractal, that every apparent power law defines a fractal dimension, or that sampling resolution can be ignored.

### C. Event-Based and Turning-Point ECG Representations

A separate line of research has explored sparse and nonuniform ECG representations. Event-driven sampling, level-crossing methods, and feature-oriented encodings replace uniformly spaced samples with events selected by the signal, usually to reduce transmission, storage, or computational cost. More recently, ECG self-similarity has been used to reconstruct morphology from event-based samples, illustrating both the efficiency of selective sampling and the risk of losing clinically meaningful waveform detail when the event set becomes too sparse [10].

Turning vertices provide a direct event definition for a sampled trajectory. Samples that continue the same direction of change belong to a monotonic run, whose accumulated amplitude excursion can be represented by its endpoints. When the direction reverses, a new vertex is created. The retained samples are therefore local extrema in the discrete, sampled-signal sense; they should not be called continuous-domain inflection points, which are defined through changes in curvature and require a different mathematical construction.

Most sparse ECG methods are evaluated through compression ratio, reconstruction error, detection performance, or energy consumption. Those objectives are important, but they do not establish that the retained-event pattern is itself a useful dynamical object. A method can reconstruct a waveform accurately while saying little about how event density, excursion magnitude, omitted continuity, or scale persistence changes from one cardiac cycle to the next. The present work takes that changing organization, rather than reconstruction efficiency, as its primary object of study.

### D. From Signal Reduction to a Dynamical Observation Domain

Against this background, FoxTail asks what becomes observable when monotonic continuity is recorded implicitly and changes of direction provide the coordinate system. The retained amplitudes describe the vertices reached by each cycle, their successive differentials describe the excursions between those vertices, and omission lengths quantify how much sampled continuity lies between events. R–R duration and original sample positions remain available as parallel descriptors rather than being absorbed into a normalized cardiac phase.

FoxTail therefore occupies a middle ground between direct waveform inspection and nonlinear summaries of the signal. Its sample-selection rule resembles event-based encoding, but its purpose is not to reconstruct the original trace from fewer samples. By superimposing the retained event sequences from many cardiac cycles, FoxTail makes beat-to-beat dispersion and changes in their organization directly visible, while allowing analysis to be repeated at different smoothing scales. Instead of reducing scale dependence to a single complexity value, the resulting descriptors remain tied to event patterns that can still be examined in the individual cycles.

The remaining gap is therefore not a lack of ECG compression methods, R-peak detectors, or nonlinear cardiac indices, but rather the absence of a representation designed specifically to expose how the internal directional structure of the ECG changes across large populations of cardiac cycles while retaining separate access to amplitude, omitted continuity, R– R timing, and scale dependence.

## III. The FoxTail Representation

### A. Computational pipeline

The representation of FoxTail was originally thought to be dynamically displayed on a monitor, along with canonical ECG traces. The processing of the raw ECG signal is described in this section, using data files from the Physionet databases [11], better described ahead.

This pipeline comprehends: reading the data and metadata from each file →identifying *or* calculating the R peaks and the R–R cycle → monotonic sample suppression →creating of event differentials →updating the window descriptors, the multiscale analysis and the statistical analyzes.

### B. R-Peak-Anchored Cardiac Cycles

Let *x*[*n*] be a discrete ECG signal acquired at sampling frequency *f*_*s*_ samples/s, and let *r*_*i*_ and *r*_*i*+1_ be consecutive R-peak landmarks. We denote the complete, uniformly sampled *i*th R-peak-anchored cardiac cycle by

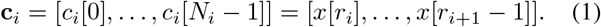

Thus, *c*_*i*_[*m*] = *x*[*r*_*i*_ + *m*] for 0 ≤ *m < N*_*i*_. The cycle includes the first anchor and excludes the next, and its original sample count and R–R duration are

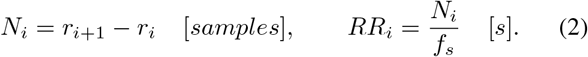

No stretching, compression, or resampling to a normalized cardiac phase is performed at this stage, so natural R–R variability remains explicit through *N*_*i*_ and *RR*_*i*_.

### C. Turning-Vertex Selection

We proceed to process *c*_*i*_ to remove monotonic intermediate samples. Define the first difference within a cycle as

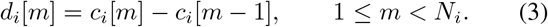

The algorithm follows the sign of successive non-zero differences. As long as that sign is unchanged, the trajectory continues along the same monotonic rise or fall and its intermediate samples are omitted from the event representation. A change of sign marks the end of one monotonic run and the beginning of another, so the shared endpoint is retained as a turning vertex. The first and last samples are retained as boundary vertices in every cycle.

Equal adjacent samples require an explicit convention because quantization can create short plateaus. We treat zero differences as continuation rather than as new events; when a plateau is followed by a reversal, its final sample represents the turning vertex. With this convention, plateaus do not become artificial clusters of retained events. The retained sample *indices* are

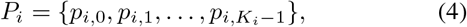

where *p*_*i*,0_ = 0, *p*_*i,Ki*_−1 = *N*_*i*_ − 1, and *K*_*i*_ is the number of retained vertices. The reduced event sequence is then defined separately from the original cycle as

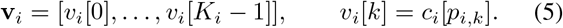

Throughout the paper, **c**_*i*_ therefore refers only to the complete sampled cycle, whereas **v**_*i*_ refers to the shorter sequence obtained after monotonic-run suppression. Its elements are discrete turning vertices or local extrema, not continuous-domain inflection points, which are defined through changes in curvature.

Fig. 1 follows the transformation from a conventional ECG to the retained vertices of one cycle and then to the amplitude and differential ensembles. Gray denotes the conventional sampled signal, red marks the R anchors and differential cycles, and blue marks retained vertices and amplitude cycles. In the ensemble panels, the black trace is the pointwise median across all cycles that still contain an event at that horizontal position. The horizontal coordinate in those panels is the event index *k*; it does not represent elapsed time, original sample position, or normalized cardiac phase.

**Fig. 1.**
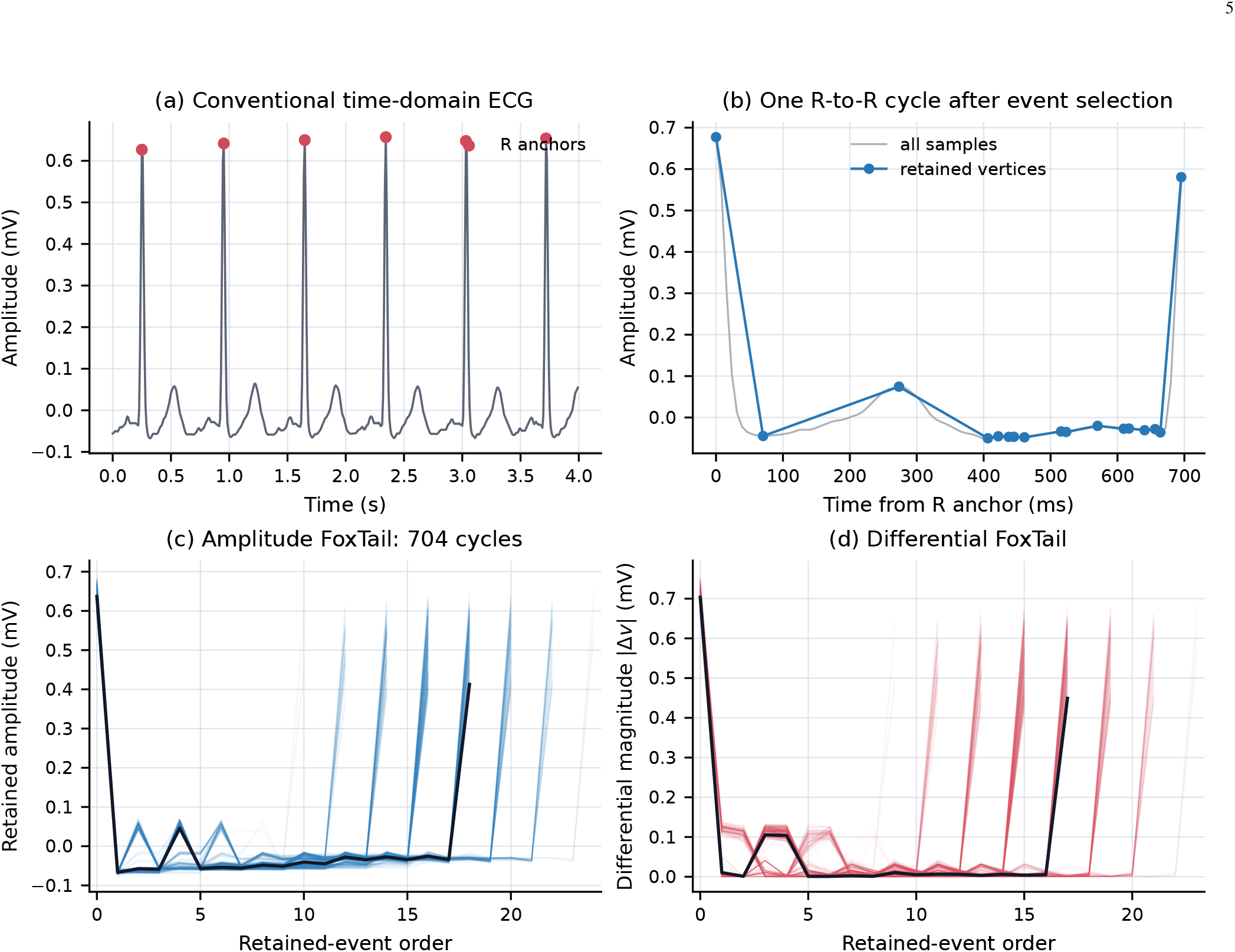
Construction of FoxTail. (a) The conventional ECG is shown in gray and the locally refined R anchors in red. (b) For one R-to-R cycle, all samples are light gray and the retained turning vertices are blue. (c) The amplitude ensemble contains one translucent blue trace per cycle. (d) The differential-magnitude ensemble contains one translucent red trace per cycle. In (c) and (d), black is the pointwise median while at least 25% of cycles still contribute an event. The example uses AFPDB n01; annotation fiducials were shifted to the local positive R maximum within ± 80 ms. Late high vertices arise when the compulsory final sample lies on the upstroke immediately before the next, excluded R peak.

### D. Two Complementary FoxTail Views

The amplitude view overlays the reduced sequences *v*_*i*_[*k*] without normalizing their vertical scale. In this form, the R-anchor amplitude and the amplitudes of all subsequent vertices remain visible, making the display sensitive to beat-to-beat morphology as well as to electrode geometry and lead orientation. Because it preserves a direct visual relationship with the conventional ECG, this is the most immediately interpretable form of FoxTail.

For analysis, consecutive vertices define signed differentials and their magnitudes:

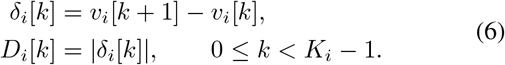

A constant baseline offset cancels in *δ*_*i*_[*k*], while its sign records the direction of the excursion and *D*_*i*_[*k*] records its magnitude. The magnitude representation produces the characteristic positive tail display used in several analyses below. The retained sequence can be recovered from *v*_*i*_[0] and the signed differentials by cumulative summation, although neither view reconstructs the samples omitted inside each monotonic run unless their original locations and values have been stored separately.

### E. Retained and Omitted Information

For each cycle, the event count and retention ratio are

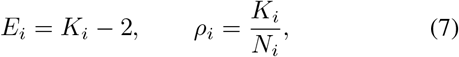

where *E*_*i*_ excludes the two compulsory endpoints. The omitted-point count is *O*_*i*_ = *N*_*i*_ −*K*_*i*_. Between two consecutive retained vertices, the omitted monotonic-run length is

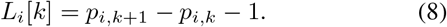

The two quantities describe complementary aspects of the same run: *D*_*i*_[*k*] measures how far the signal travels between vertices, whereas *L*_*i*_[*k*] records how many uniformly sampled points were omitted along that excursion. Across beats, the standard deviation, SD(*ρ*_*i*_), describes dispersion in event density. The related quantity SD(Δ*ρ*_*i*_), with Δ*ρ*_*i*_ = *ρ*_*i*+1_ − *ρ*_*i*_, describes how abruptly the retention ratio changes from one cycle to the next. They must be interpreted jointly because the same retention ratio may arise from stable physiology, evolving morphology, high-frequency contamination, low-frequency movement, sampling resolution, or anchor error.

### F. A Supporting Mathematical Guarantee

The algorithm discards intermediate samples, but not the accumulated absolute variation along a monotonic run. Since the absolute first differences telescope between consecutive retained vertices, the Total Variation for the *i*^*th*^ cycle is

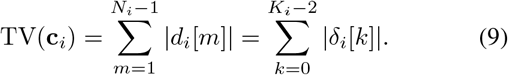

Consequently, the discrete total variation is preserved exactly, apart from floating-point round-off errors. This identity guarantees one property of the transformation, but it should not be read as a claim that all ECG information is preserved: uniform timing inside runs, curvature, signal energy, and the complete intermediate morphology are deliberately absent from **v**_*i*_.

### G. Multiscale Event Persistence

Let *x*_*σ*_[*n*] denote the ECG after Gaussian smoothing with scale parameter *σ*, defined as the Gaussian standard deviation in milliseconds; *σ* = 0 denotes the unsmoothed reference signal. Applying the same anchors and turning-vertex rule at each scale produces *E*_*i*_(*σ*) and *ρ*_*i*_(*σ*), allowing events that disappear after minimal smoothing to be distinguished from those that persist as progressively finer structure is removed. We selected *σ* ∈ {0, 8, 16, 32, 64, 128} ms for three related reasons. First, after harmonization to 128 Hz, 8 ms is approximately one sampling interval, so the positive scales correspond closely to Gaussian standard deviations of 1, 2, 4, 8, and 16 samples. Second, this dyadic spacing samples the scale axis approximately uniformly in log_2_ *σ*, which supports comparisons of persistence and avoids assigning disproportionate weight to a narrow band of fine scales. Third, the range extends from sample-scale perturbations to a coarse within-cycle representation, while its upper bound remains below a typical adult R–R interval. These values were chosen as an exploratory, computationally transparent scale set rather than as exact boundaries between physiological ECG components; in short tachyarrhythmic cycles, 128 ms may occupy a substantial fraction of the cycle.

When the record-level median interior event count follows

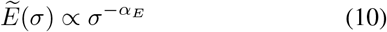

over 8–128 ms, *α*_*E*_ is estimated by log–log regression. We use it only as an empirical descriptor of how rapidly the median event count falls with smoothing. Interpreting it as a fractal dimension would require a wider and independently justified scaling range, evidence of estimator stability, and a formal correspondence between the retained-event process and an appropriate fractal model.

## IV. Experimental Design

### A. Databases and Scientific Roles

The evaluation used five open PhysioNet resources [11], [12]. Four provided distinct physiological contexts for visualization and quantitative description, while NSTDB supplied a controlled measurement-noise experiment. Table I distinguishes their record counts, subjects or record sets, acquisition characteristics, and scientific roles. All successfully processed first-channel records were included, with the single exception of CUDB record cu21, whose annotation file yielded fewer than two beat anchors; consequently, 34 of the 35 CUDB records remained available for analysis.

**TABLE 1.** Databases and Their Roles in the foxTail Evaluation.

| Database | Records | Native $f_s$ | Population or signal context | Role in the scientific argument |
| --- | --- | --- | --- | --- |
| AFPDB [13] | 300 | 128 Hz | Paired 30-min records and 5-min continuations from PAF, non-documented-AF, and blinded challenge sets | Primary within-subject test: 25 PAF record sets comparing the continuation after the PAF-distant excerpt with the continuation containing the transition from the pre-PAF excerpt into PAF. |
| NSRDB | 18 | 128 Hz | Long-term recordings without significant arrhythmia | Stable-rhythm visual and descriptive reference; not a control group for causal cross-database inference. |
| CHFDB [14] | 15 | 250 Hz | Approximately 20-h recordings from subjects with severe NYHA III–IV heart failure | Chronic pathological context and visual stress test of long-record representation. |
| CUDB [15] | 34 | 250 Hz | Short recordings containing sustained ventricular tachycardia, flutter, or fibrillation | Acute-transition test using windows before the first approximate VF-onset annotation. |
| NSTDB [16] | 12 | 360 Hz | Two clean source ECGs contaminated with calibrated electrode-motion noise at six SNR levels | Controlled test of whether event density and related descriptors behave as simple proxies for noise. |

The AFPDB learning set contains paired 30-min excerpts from the same long-term ECG. For PAF subjects, the odd-numbered record is distant from PAF and the even-numbered record immediately precedes an episode; their respective five-minute continuations confirm the absence or presence of the transition [13]. Because the supplied `qrs` annotations are machine generated and unaudited, we treated them as reference fiducials rather than as manual ground truth. A similar caution applies to the automated, uncorrected CHFDB beat annotations and to CUDB, where VF boundaries are approximate and all beats are nominally labelled normal despite frequent ectopy. These properties of the source databases necessarily limit the physiological specificity of the analysis.

The most controlled comparison is provided by NSTDB, in which electrode-motion artifact was added to clean MIT-BIH records 118 and 119 at SNRs of −6, 0, 6, 12, 18, and 24 dB. Beginning after the first five minutes, two-minute contaminated and clean intervals alternate while the original beat annotations remain available [16]. This construction separates signal identity from noise level, although the twelve records are derived from only two independent source ECGs.

### B. Signal Preparation and Anchoring

The WaveForm DataBase (WFDB) software and file format provide standardized access to PhysioNet waveform samples, header metadata, channel gains, and annotations [12]. Using the gain and analog-to-digital converter offset recorded in each

WFDB header, digital samples were converted to physical units as

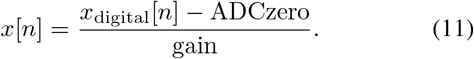

Non-finite values were interpolated temporarily to permit filtering, but every cardiac cycle containing an originally invalid sample was subsequently excluded. The numerical batch used the first available ECG channel and retained its physical amplitude without normalization.

We computed both native and harmonized representations. Native mode retained each database’s original sampling frequency, whereas harmonized mode applied a fourth-order zero-phase Butterworth bandpass from 0.5 to 40 Hz and then resampled the signal to 128 Hz by polyphase filtering. Whenever the native Nyquist limit was lower, the high cutoff was restricted to 95% of Nyquist. Reference annotations were mapped to the resulting sampling grid and used directly as QRS anchors in the numerical batch.

The illustrative ensembles in Figs. 1–3 required a stricter visual definition of the anchor. For these figures, each annotation was refined to the local positive R maximum within ±80 ms. We displayed the second channel for AFPDB n07 and CHFDB chf03, the first channel for NSRDB 17453, and the only channel for CUDB cu09. The CUDB trace was multiplied by −1 because its dominant QRS deflection was negative. This whole-record polarity change does not normalize amplitude; it only makes the displayed R landmark upright and prevents an annotation fiducial immediately before the R maximum from appearing as event zero. Importantly, an R peak is a local QRS landmark, not a guarantee that no later deflection or artifact in the cycle has greater amplitude.

The primary analyses use harmonized mode so that the probability of observing a turning point is compared on a common temporal grid. Native-mode results remain in the analysis output as a sensitivity representation. Harmonization reduces, but does not eliminate effects resulting from lead, hardware, annotation, and database changes. Cross-database contrasts are therefore descriptive and are not interpreted as disease effects.

### C. Windows and Record-Level Summaries

Descriptors were calculated for complete R-to-R cycles and summarized over the full record, in non-overlapping five-minute windows, and in 60-s windows advanced every 30 s. Only windows containing at least 30 complete cycles were retained. Cycle-level quantities were summarized by their median and interquartile range, *D*_*i*_[*k*] and *L*_*i*_[*k*] were pooled across all events within the window, and SD(*ρ*_*i*_) and SD(Δ*ρ*_*i*_) were evaluated in beat order.

Gaussian smoothing was applied at *σ* = 0, 8, 16, 32, 64, and 128 ms while the anchors remained fixed. Scaling exponents were fitted to record-level median *E*_*i*_(*σ*) at 8–128 ms when at least four positive scale values were available.

### D. Prespecified Quantitative Questions

The experiments were organized around four questions that follow directly from the proposed purpose of the representation.

**First**, what does FoxTail make visible across distinct ECG contexts? Representative records were selected before inspection of their images as the record closest to the group median harmonized retention ratio. AFPDB n07 was selected within the full, non-documented-AF recordings; NSRDB 17453, CHFDB chf03, and CUDB cu09 were selected within their respective databases. Figures use the first 600 s, except for CUDB cu09, which was truncated at its first VF annotation after 239.1 s.

**Second**, does the event domain detect a change within the same subject? For each of the 25 AFPDB PAF record sets, the five-minute PAF-onset continuation of the even, pre-PAF record was compared with the continuation of the odd, PAF-distant record. The three primary descriptors were median *ρ*, SD(Δ*ρ*), and *α*_*E*_.

**Third**, does controlled noise produce a simple increase in turning-point density? Harmonized record-level descriptors were followed across the six NSTDB SNR levels separately for source records 118 and 119.

**Fourth**, does the event domain show a common short-term transition before VF? For the 29 CUDB records with a VF-onset annotation and sufficient paired windows, per-record medians from the final two minutes were compared with those from two to six minutes before VF. The CUDB documentation states that its VF-onset annotations are approximate and that the recorded fibrillation episodes are generally preceded by ventricular tachycardia [17]. The comparison is therefore a stringent test for a shared final transition, not a contrast between normal rhythm and ventricular arrhythmia.

### E. Statistical Analysis

The inferential unit was the subject or record rather than the cardiac cycle, thereby avoiding the pseudoreplication that would result from treating thousands of beats as independent observations. Paired differences were summarized by their median, and confidence intervals for the AFPDB medians were obtained from 20,000 subject-level bootstrap resamples using a fixed random seed. Two-sided Wilcoxon signed-rank tests assessed paired changes, with Holm correction across the three primary AFPDB metrics and, separately, across the five exploratory CUDB descriptors. NSTDB was analyzed descriptively because its twelve noise-stress records arise from only two independent source ECGs. Since the purpose was to evaluate an observation domain rather than diagnostic discrimination, no disease classifier was trained.

## V. Results

### A. What a FoxTail Looks Like

Figures 2 and 3 present the same four representative records in complementary forms. Blue denotes AFPDB, green NSRDB, purple CHFDB, and orange CUDB. Within each panel, every translucent colored line is one cardiac cycle; overlapping cycles create darker regions. The black trace is not an additional beat: it is the pointwise median while at least 25% of the cycles still contain an event at that position. The amplitude view preserves the locally refined R-anchor amplitude and the subsequent retained trajectory, whereas the differential-magnitude view removes constant baseline offsets and emphasizes excursion size. Both displays preserve variable sequence length and use event order, rather than time, as their horizontal coordinate.

**Fig. 2.**
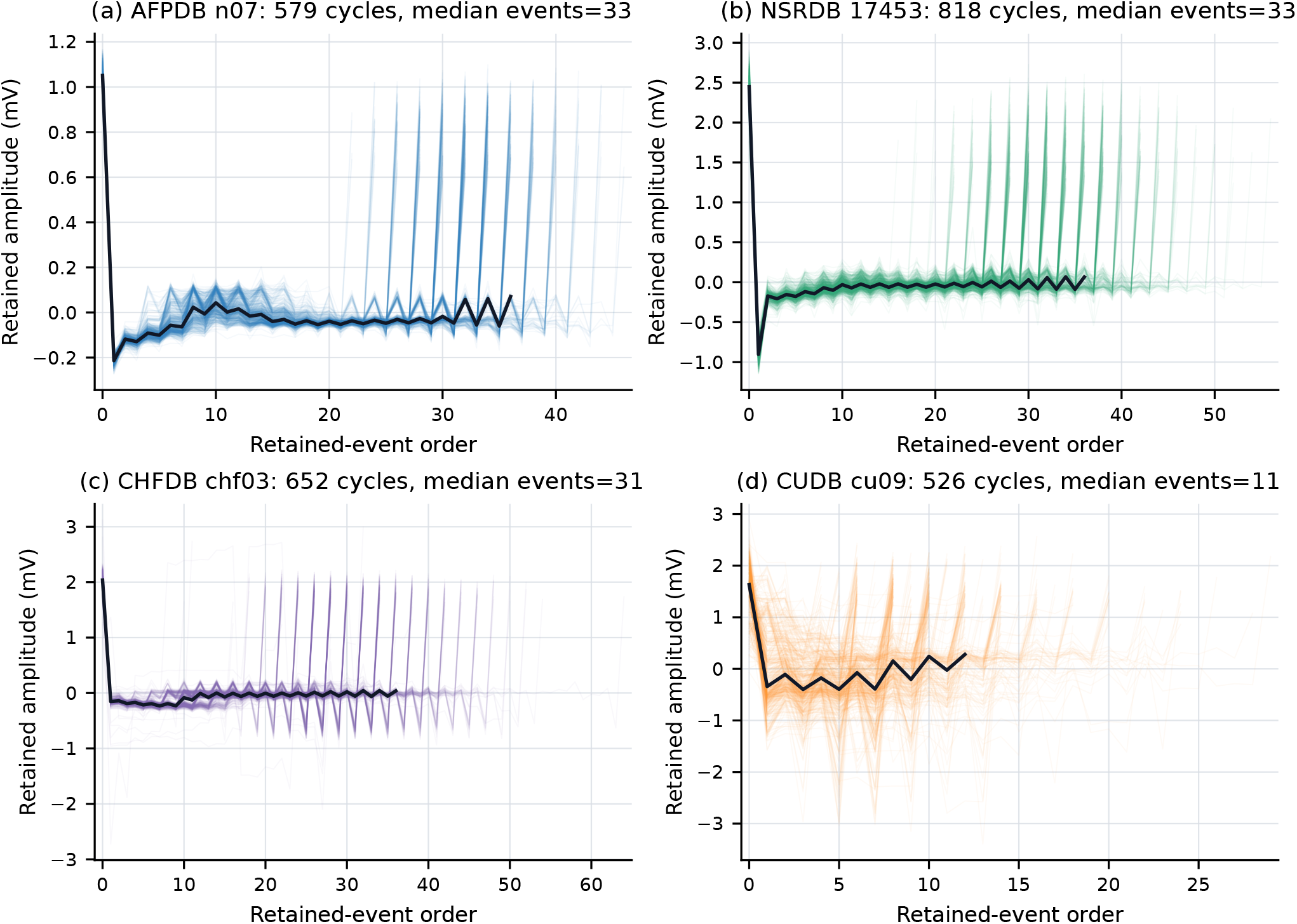
Representative amplitude FoxTail ensembles after harmonization and local R-peak refinement. Blue, green, purple, and orange identify AFPDB, NSRDB, CHFDB, and CUDB, respectively. Each translucent colored trace is one complete R-to-R cycle, and black is the pointwise median while at least 25% of cycles remain. The displayed channel was chosen to provide an upright dominant QRS complex; the sole CUDB channel was polarity-inverted. Panel-specific axes remain in mV and no amplitude normalization was applied. Late high vertices can be the compulsory sample immediately before the next, excluded R peak. These examples show the visual range of the representation, not diagnostic signatures.

**Fig. 3.**
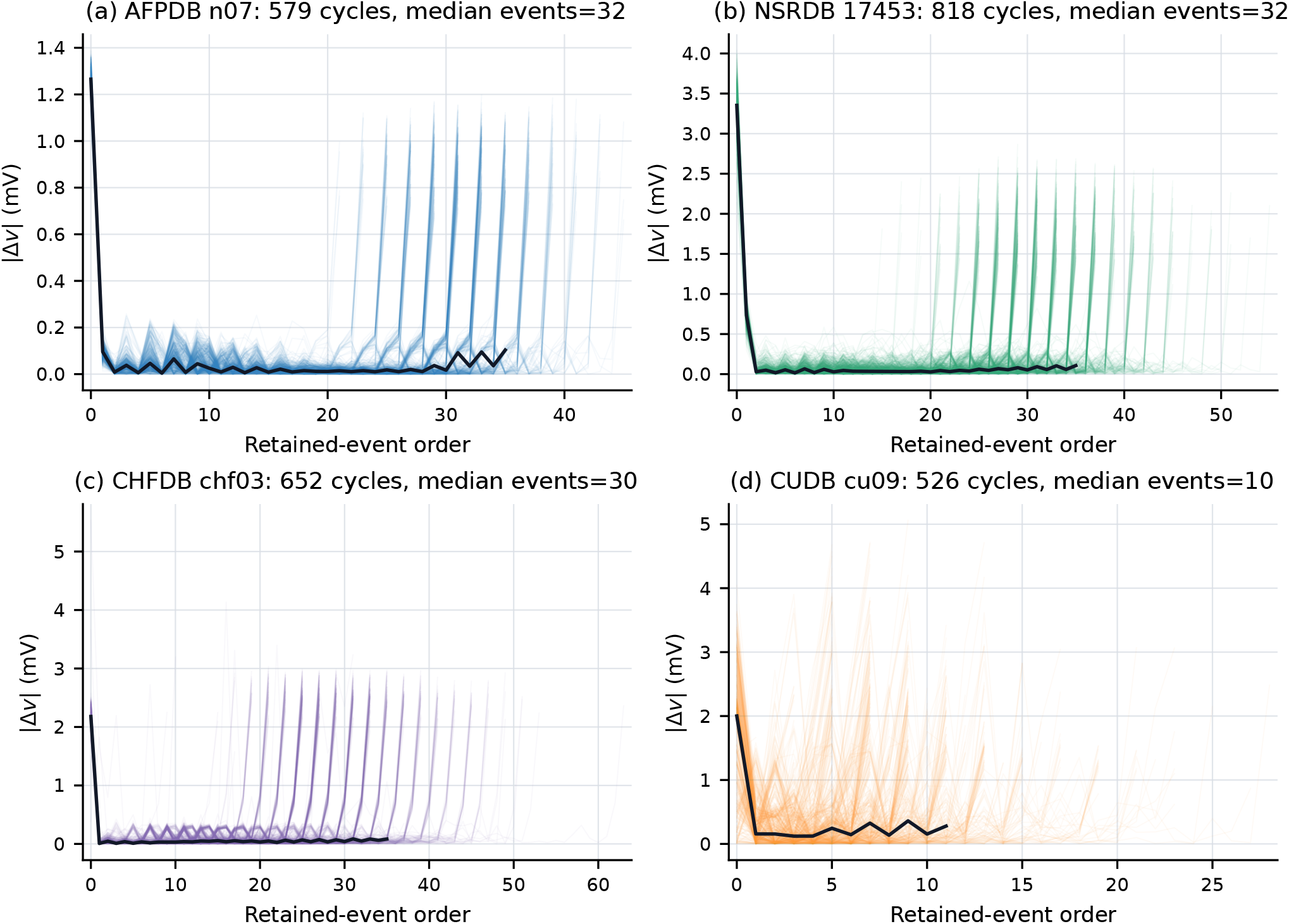
Differential-magnitude FoxTail ensembles for the same records and cycles as Fig. 2. Database colors and the black pointwise-median trace have the same meaning. The vertical coordinate is |Δ*v*| between consecutive retained vertices. Event density, excursion magnitude, dispersion, and variable tail length are simultaneously visible. These are examples of the observational domain, not evidence of diagnostic separation.

The four ensembles differed visibly in their proximal organization, excursion size, dispersion, and tail length. Within the displayed intervals, AFPDB n07 contained 579 cycles with a median of 33 retained vertices (32 differentials), NSRDB 17453 contained 818 cycles with 33 vertices (32 differentials), CHFDB chf03 contained 652 cycles with 31 vertices (30 differentials), and the pre-VF portion of CUDB cu09 contained 526 cycles with 11 vertices (10 differentials). The repeated distal spikes in some panels occur because cycles of different event counts reach the compulsory pre-next-R endpoint at different horizontal positions. These counts and shapes should not be interpreted as an ordering of clinical complexity; they show that different combinations of rate, morphology, measurement, and physiological state can produce different event-domain organizations.

### B. Descriptor Landscapes Across Databases

Table II summarizes record-level harmonized descriptors before smoothing (*σ* = 0). Each column answers a different question. Median *ρ* is the typical fraction of uniformly sampled points retained as vertices; SD(Δ*ρ*) measures how irregularly that fraction changes from one beat to the next; median *D* is the typical amplitude excursion between consecutive vertices; and median *L* is the typical number of samples omitted along a monotonic run. The table reports the median and interquartile range of each record-level summary across a database, so the entries describe between-record landscapes rather than a pooled population of beats.

**TABLE 2.** Record-Level foxTail Descriptor Landscapes in Harmonized Mode. Values Are Median [Interquartile Range] Across Records.

| Database | Records | Median $\rho$ | SD($\Delta\rho$) | Median $D$ (mV) | Median $L$ |
| --- | --- | --- | --- | --- | --- |
| AFPDB | 300 | 0.3114 [0.2774, 0.3465] | 0.0495 [0.0444, 0.0560] | 0.0224 [0.0159, 0.0394] | 1 [1, 1] |
| NSRDB | 18 | 0.3211 [0.2922, 0.3405] | 0.0492 [0.0461, 0.0522] | 0.0252 [0.0213, 0.0298] | 1 [1, 1] |
| CHFDB | 15 | 0.2479 [0.2254, 0.2843] | 0.0511 [0.0455, 0.0565] | 0.0253 [0.0205, 0.0598] | 2 [1, 2] |
| CUDB | 34 | 0.1979 [0.1595, 0.2460] | 0.0545 [0.0434, 0.0639] | 0.0708 [0.0421, 0.1323] | 2 [1.25, 3] |
| NSTDB | 12 | 0.3133 [0.3096, 0.3226] | 0.0497 [0.0484, 0.0508] | 0.0565 [0.0417, 0.0804] | 1 [1, 1] |

NSRDB and AFPDB retained approximately one sample in three, with median *ρ* values of 0.3211 and 0.3114. CHFDB retained about one in four (0.2479), while CUDB retained about one in five (0.1979). The CUDB records nevertheless had the largest typical differential magnitude (0.0708 mV) and longer omitted runs, showing why low retention cannot be equated with low signal activity. Beat-to-beat instability was comparatively similar across databases, with median SD(Δ*ρ*) between 0.0492 and 0.0545. After smoothing, the database profiles converged in Fig. 5a: at *σ* = 128 ms, median retention ratios ranged only from approximately 0.039 to 0.049. Most of the visible separation in event density therefore resides at finer temporal scales.

These cross-database differences are presented descriptively because disease state is collinear with database identity for NSRDB, CHFDB, and CUDB. Consequently, neither Table II nor the visual examples can separate physiology from lead placement, recording hardware, annotation practice, or population composition.

### C. Within-Subject Change Around PAF

The paired AFPDB analysis tested within-subject change without relying on cross-database separation. In Table III, every difference is defined as PAF-onset continuation minus PAF-distant continuation. The negative change in *ρ* therefore means that the onset-associated continuation retained slightly fewer vertices, whereas the positive change in SD(Δ*ρ*) means that its event density varied more abruptly from beat to beat. The negative change in *α*_*E*_ requires a different interpretation: under Eq. (10), a smaller exponent describes a shallower decline in event count as smoothing increases, and thus greater persistence of retained structure across the tested scales. All three rank-based paired tests remained significant after Holm correction.

**TABLE 3.** Paired AFPDB Changes: PAF-Onset Continuation Minus PAF-Distant Continuation (*n* = 25 SUBJECTS).

| Descriptor | Median paired difference | Bootstrap 95% CI | Wilcoxon $p$ | Holm $p$ |
| --- | --- | --- | --- | --- |
| Median retention ratio $\rho$ | -0.00768 | $[-0.02634, -0.00378]$ | 0.02298 | 0.02298 |
| $SD(\Delta\rho)$ | +0.01016 | $[+0.00624, +0.01416]$ | 0.000912 | 0.002736 |
| Scaling exponent $\alpha_E$ | -0.05850 | $[-0.17078, 0.00000]$ | 0.004925 | 0.009850 |

The bootstrap interval for the median *α*_*E*_ difference reaches zero at the reported numerical precision, even though the Wilcoxon test remains significant. These statements are not contradictory: the interval estimates the median difference, whereas the Wilcoxon test uses the signed-rank distribution of all paired differences. Figure 4 makes that distribution visible. Gray lines connect the two continuations from each subject, blue points show the distant condition, red points the onset-associated condition, and black diamonds join the two group medians. Individual trajectories vary, but their aggregate direction agrees with the table.

**Fig. 4.**
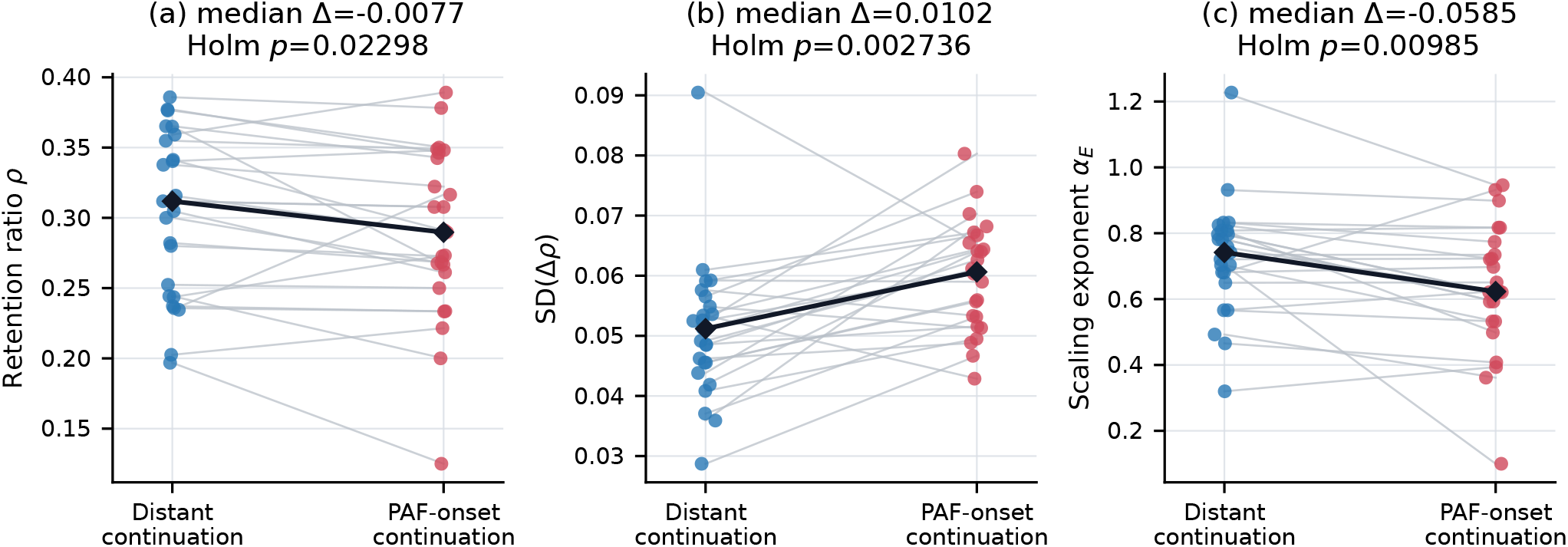
Within-subject AFPDB comparison. Gray lines join the PAF-distant continuation (blue) and PAF-onset continuation (red) for each subject; black diamonds show group medians. Panel (a) shows slightly lower typical retention, (b) greater successive retention instability, and (c) a lower multiscale exponent in the onset-associated continuation.

### D. Noise Is Not Equivalent to Event Maximization

Figure 5 combines the scale and noise experiments. In panel (a), each colored curve is the database median and its shaded region is the interquartile range across records. All curves fall as smoothing removes fine directional reversals, but their separation is greatest at the smallest scales and narrows substantially by *σ* = 128 ms. Panels (b)–(d) follow the two independent NSTDB source ECGs separately: red identifies record 118 and blue record 119, with each point corresponding to one calibrated signal-to-noise ratio (SNR).

**Fig. 5.**
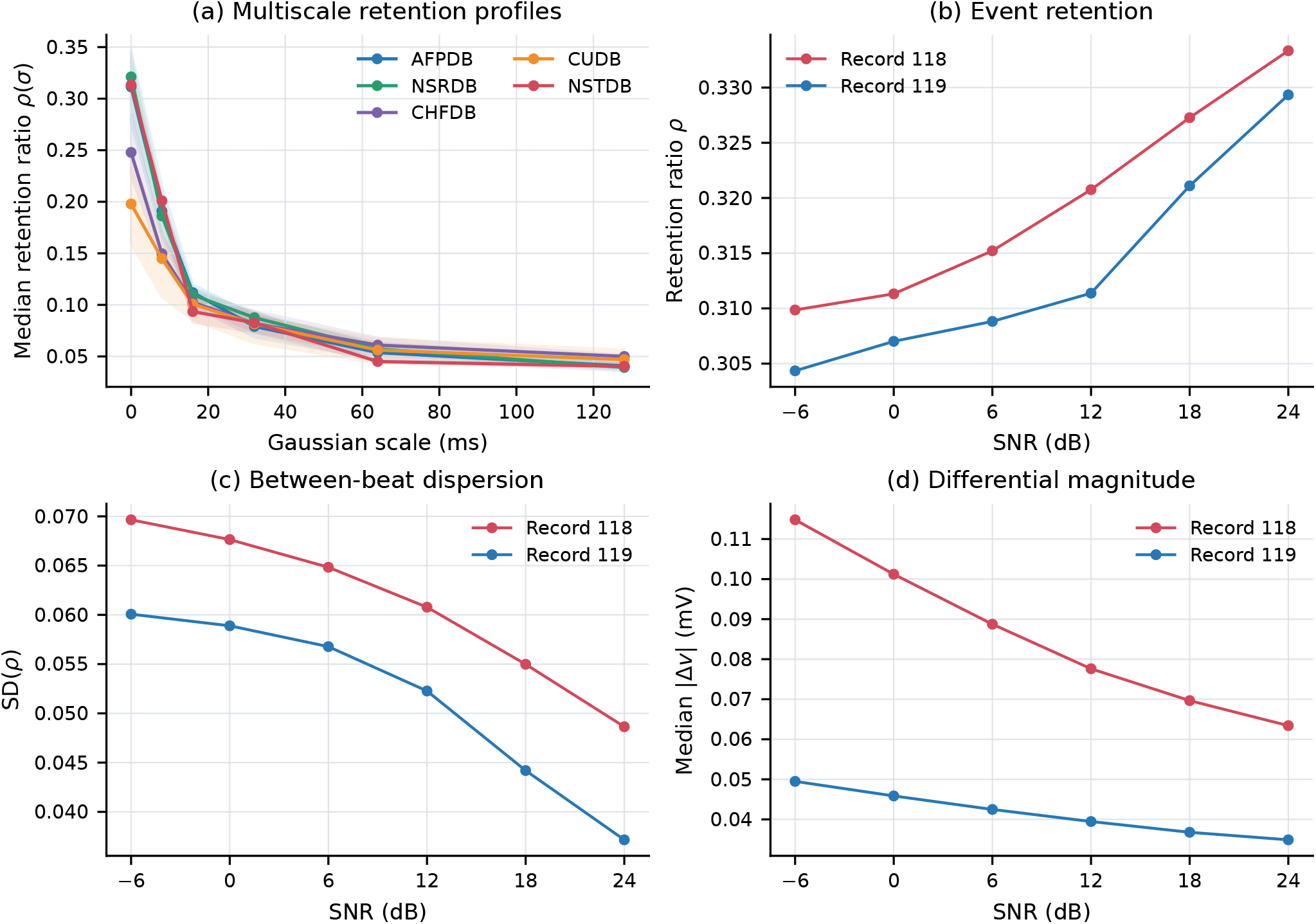
Scale and noise sensitivity. (a) Database medians and interquartile bands: AFPDB blue, NSRDB green, CHFDB purple, CUDB orange, and NSTDB red. (b)–(d) Harmonized NSTDB descriptors across calibrated electrode-motion noise levels; record 118 is red and record 119 blue. Lower SNR reduced typical retention but increased between-beat dispersion and differential magnitude. Realistic movement artifact therefore did not behave like a homogeneous high-frequency perturbation.

The NSTDB experiment contradicted the simplest expectation that increasing noise must maximize the number of turning events. As SNR increased from −6 to 24 dB, meaning that less noise was present, the mean harmonized retention ratio rose monotonically from 0.3071 to 0.3313. Over the same range, mean SD(*ρ*) fell from 0.0649 to 0.0429 and median differential magnitude fell from 0.0822 to 0.0492 mV (Fig. 5b–d). The same directional pattern appeared in both source ECGs, although their absolute values differed.

This result is consistent with the character of NSTDB electrode-motion artifact, which is transient and can create large, slowly varying baseline excursions and beat-like disturbances rather than a homogeneous sequence of high-frequency sign changes. The proposition that noise necessarily increases the number of turning points is therefore too simple: a credible noise interpretation must consider retention together with variability, amplitude, temporal localization, and scale persistence.

### E. No Common Final Pre-VF Signature in CUDB

Among 29 CUDB records with sufficient paired windows, no primary event-domain descriptor changed robustly in the final two minutes relative to two to six minutes before the annotated VF onset. Median paired changes were −0.00269 for *ρ*, −0.00156 for SD(Δ*ρ*), and +0.00314 mV for median *D*, with Holm-adjusted *p* = 0.288, 1.000, and 0.593, respectively. Median *L* had an unadjusted *p* = 0.0361 but did not survive correction (*p* = 0.180).

Figure 6 shows why the group statistics are small. Gray subject-level lines move in both directions between the earlier window (blue) and final window (orange), while the black median diamonds remain close together. The figure therefore contains substantial individual change but no consistent database-wide direction. That heterogeneity is scientifically different from an absence of variation: it argues against a single final two-minute signature while leaving open earlier or subject-specific transitions.

**Fig. 6.**
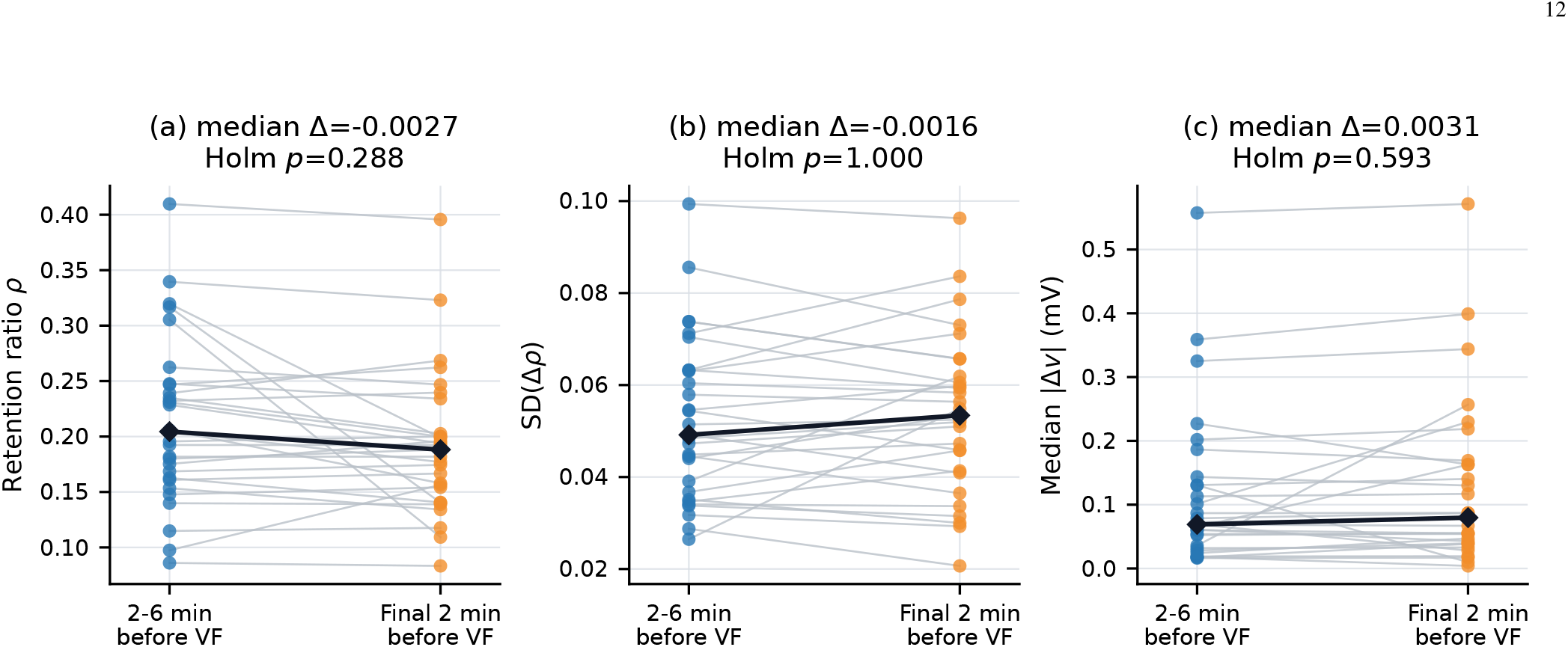
CUDB temporal test. Gray lines connect each record’s earlier window (blue; two to six minutes before VF) to its final window (orange; final two minutes); black diamonds show group medians. The mixed line directions and nearly unchanged medians explain why no displayed descriptor survived multiplicity correction.

The absence of a common final transition does not show that the representation is insensitive to ventricular tachyarrhythmia. CUDB recordings often contain sustained tachycardia well before the annotated boundary, the onset of fibrillation is approximate, and the relevant change may occur earlier or differ between individuals. The result does show that an appealing representation should not be promoted as a universal precursor without a successful temporal test.

### F. Total Variation Identity

We verified Total Variation (Eq. (9)) for every analyzed cycle. The maximum relative discrepancy between sample-domain total variation and the sum of differential magnitudes was 1.01×10^*−*15^, which is consistent with floating-point round-off and confirms the numerical implementation of the monotonic-run reduction. Because this verification does not alter the empirical findings, it is treated as a validation check rather than as the central result.

## VI. Discussion

### A. The Main Finding Is a New Observation Domain

The central result is not that an ECG can be compressed or that one scalar quantity can be preserved. Rather, cardiac cycles can be reorganized into an R-anchored event domain in which variation across hundreds of beats remains visible and gives rise to interpretable measurements. The amplitude view preserves anchor and vertex variability for human inspection, while the differential view separates successive excursions from constant baseline offset. In both cases, event counts, retention, omitted continuity, instability, and persistence across scale remain connected to the visible ensemble instead of appearing as detached numerical features.

The paired AFPDB result provides the strongest present evidence that this domain carries state-related information. Although acquisition system and subject identity were held constant, the PAF-onset continuation associated with the pre-PAF recording differed from the distant continuation in three complementary ways: typical retention decreased slightly, successive variability increased, and the multiscale event-count profile declined more gradually. Describing this merely as “more complexity” would obscure the structure of the result, because the joint pattern is more informative than the direction of any single descriptor.

### B. Why Preservation Is Secondary

Exact total-variation preservation answers a necessary objection by showing that monotonic-run suppression is not an arbitrary decimation of accumulated excursion. It does not amount to general information preservation, because the transform deliberately exchanges a uniformly sampled temporal trajectory for an ordered event trajectory. Timing remains available only through *RR*_*i*_, retained indices, or *L*_*i*_[*k*], while curvature, signal energy, and the precise waveform inside a monotonic run cannot be recovered from **v**_*i*_ alone. The value of FoxTail therefore depends on whether this trade makes relevant changes observable, which is why the paired, noise, and temporal experiments carry more scientific weight than the identity itself.

### C. Noise, Complexity, and Scale

Turning-point density is sensitive to noise, but both the direction and the form of that sensitivity depend on the disturbance. Homogeneous high-frequency perturbations may increase sign reversals, whereas NSTDB electrode-motion artifact reduced typical retention after harmonization while increasing between-beat dispersion and excursion magnitude. Because movement artifact is temporally localized and spectrally structured, it can deform entire cycles rather than merely add small reversals. Noise detection should therefore rely on multivariate and temporal patterns, including whether changes are persistent, intermittent, localized, and scale dependent.

The multiscale results must also be interpreted cautiously. ECG and heartbeat dynamics can exhibit fractal or multifractal organization [2], [7], [9], but self-similarity neither makes sampling frequency irrelevant nor converts *α*_*E*_ automatically into a fractal dimension. In the present study, *α*_*E*_ measures empirically how quickly retained turning events disappear under Gaussian smoothing. Establishing a formal relationship with fractal geometry will require simulations with known scaling laws, broader scale ranges, estimator comparisons, and explicit tests against non-fractal stochastic processes.

### D. Human Interpretation and Physiological Localization

The two visual forms should be evaluated by their intended users. Cardiologists may prefer absolute retained amplitudes because they maintain a more direct relationship with the conventional waveform, including R-peak amplitude variability. Engineers may prefer signed differentials or magnitudes because they remove constant offsets and expose event excursions. A reader study should compare these views for recognition of within-record changes rather than ask clinicians to diagnose directly from FoxTail.

The present transform treats the cardiac cycle as one sequence. A natural extension is to partition events into physiological regions associated with P, QRS, ST, and T activity and to analyze monotonicity, amplitude, and scale persistence within each region. This could reveal whether a global change arises predominantly from atrial depolarization, ventricular activation, repolarization, or the intervals between them. Alternative anchors at P, QRS onset, T peak, or other landmarks may also expose changes that R anchoring obscures, particularly when a landmark is absent or unstable. Those variants should remain future work until clinicians determine which views are most interpretable.

### E. Limitations

This study has six principal limitations. First, most numerical anchors were supplied automated QRS fiducials rather than locally refined R maxima. The figure audit showed that this distinction is visible in an event-ordered plot, and anchor uncertainty can change both cycle boundaries and early event order. A full sensitivity analysis should therefore repeat the numerical batch with adjudicated or locally refined landmarks. Second, the numerical batch used only the first channel, although the visual examples demonstrate that lead orientation strongly affects amplitude and QRS polarity. Third, the lack of amplitude normalization preserves potentially useful variability but also preserves gain, electrode, and hardware differences. Fourth, NSRDB, CHFDB, and CUDB labels are inseparable from database identity, so cross-database differences are descriptive. Fifth, NSTDB contains only two independent source ECGs and only electrode-motion contamination in the analyzed records. Sixth, the AFPDB result is state associated but not yet a clinical validation: continuations differ in rhythm content, annotations are unaudited, and no external cohort or blinded reader study was used.

The CUDB negative result is also constrained by approximate VF annotations, pre-existing ventricular tachycardia, short recordings, and heterogeneous trajectories. Conversely, its inclusion reduces publication bias within the manuscript by showing where the current descriptors did not yield a robust common transition.

### F. Next Validation Step

The next study should adopt a prospective, longitudinal design in which consistent multi-lead ECGs are acquired from the same individuals across well-defined physiological or clinical states. It should use adjudicated landmarks and compare changes in FoxTail with conventional ECG measures, rhythm, HRV, signal quality, and relevant clinical covariates. The primary endpoints should concern sensitivity to state change and repeatability rather than diagnosis alone, and subject-specific baselines may prove more appropriate than population thresholds. Predictive classifiers should be introduced only after measurement validity has been established, using subject-level data separation and explicit tests against acquisition confounding.

A further natural extension treats the overlaid retained-vertex sequences not only as a set of scalar descriptors but as a geometric object in its own right. Persistent homology and related tools from topological data analysis provide one principled way to characterize such an object, extracting invariant structural features that are robust to the kind of noise and non-stationarity discussed above [18]. Applying this class of methods to the amplitude and differential FoxTail ensembles could in principle reveal structure that record-level scalar descriptors such as *ρ, D*, and *L* do not separately capture, including within populations that carry no rhythm-based diagnosis, where conventional analysis has little basis for expecting internal heterogeneity in the first place. This is presented here as an open methodological question motivated by the present construction, not as a validated finding, and it would require the same discipline applied throughout this study: explicit null comparisons, correction for multiplicity, and testing against confounds such as acquisition hardware and lead placement before any structural difference is interpreted as physiological.

## VII. Conclusion

FoxTail reorganizes the ECG from uniformly sampled time into an R-peak-anchored sequence of directional events. This makes the organization of change within successive cardiac cycles visible across long recordings and measurable through amplitude, differential excursion, retained-event density, omitted continuity, beat-to-beat instability, and multiscale persistence. Paired AFPDB recordings showed that the domain can detect within-subject state-related changes, while NSTDB and CUDB demonstrated why neither event density nor visual appearance can be interpreted simplistically as noise, complexity, pathology, or imminent transition. FoxTail should therefore be understood as a complementary observation and measurement domain: a way to ask how ECG dynamics change, before asking whether those changes diagnose a disease.

## Data Availability

All data produced are available online at the Physionet database website.

https://physionet.org/

## Acknowledgment

This research was funded in whole or in part by Fundação para a Ciência e a Tecnologia, I.P. (FCT; ROR: 00snfqn58), through the following grants: UID/00645/2025 (doi:10.54499/UID/00645/2025); UID/PRR/00645/2025 (doi:10.54499/UID/PRR/00645/2025); and UID/50008/2025 (doi:10.54499/UID/50008/2025). For the purpose of Open Access, the author has applied a CC BY public copyright licence to any Author Accepted Manuscript (AAM) version arising from this submission.

## Notes

### Competing Interest Statement

The authors have declared no competing interest.

### Author Declarations

This study uses data that is publicly available at Physionet Databases.

